# Slow-SPEED: protocol for three randomised trials of remotely delivered exercise to prevent Parkinson’s disease

**DOI:** 10.64898/2026.03.05.26347705

**Authors:** Thomas H. Oosterhof, Eleanor Mitchell, Alberto Ascherio, Stella Aslibekyan, Viktoria Azoidou, Kara Beasley, Yoav Ben-Shlomo, Eline Bunnik, Camille Carroll, Lana Chahine, Daniel Corcos, Jules M. Janssen Daalen, Karin D. van Dijk, Bauke W. Dijkstra, Lisanne Dommershuijsen, Ray Dorsey, Luc J. W. Evers, Rick C. Helmich, Martin Johansson, Lucy Norcliffe-Kaufmann, Jessi Keavney, Christine Klein, Matthew J Kmiecik, Thomas Kustermann, Eric A. Macklin, Kenneth Marek, Sanne K. Meles, Sebastiaan Overeem, Carl M. Philpott, Angelique Pijpers, Ronald B. Postuma, Helen M. Rowbotham, Sabine Schootemeijer, Michael A. Schwarzschild, Tanya Simuni, Michael Sommerauer, Ambra Stefani, Kenan Steidel, Marcel Verbeek, Rick van der Vliet, Nienke M. de Vries, Bart van de Warrenburg, Richard K. Wyse, Bastiaan R. Bloem, Ruth B. Schneider, Alastair J. Noyce, Sirwan K.L. Darweesh

**Author notes:** Correspondence to: Sirwan. K.L. Darweesh, Department of Neurology, Center of Expertise for Parkinson & Movement Disorders; Radboud University Medical Center, PO Box 9101, 6500 HB Nijmegen, Netherlands. These authors contributed equally to this work (Alastair J. Noyce, Sirwan K.L. Darweesh).

## Abstract

We describe the design of the first non-pharmacological prevention trials of Parkinson’s Disease worldwide: the randomised controlled ‘Slow-SPEED’ trials. The three trials examine the feasibility and preliminary efficacy of a gamified, remotely administered exercise intervention vs. active control program over 18-36 months in the Netherlands (n=110), United Kingdom (n=110) and United States (n=600). Each trial focuses on a complementary prodromal subgroup: isolated/idiopathic REM sleep behavioural disorder, hyposmia, or *LRRK2*/*GBA1* mutation carriers. These trials will provide unique insights for large-scale Parkinson’s Disease prevention studies.

## Background and rationale

### Scientific and social relevance

Parkinson’s disease (PD) affected nearly 12 million people globally in 2021, a prevalence substantially above a prediction of 7 million made as recently as 2018^1-3^. To date, no disease-modifying treatment for PD is available. PD diagnosis is based on the recognition of distinct motor signs and symptoms, classically bradykinesia, rigidity and tremor. These hallmark features are identified at a relatively advanced stage of neurodegeneration, by which time the underlying damage may be irreversible^4^. This may be one reason why disease modifying trials in those with clinically manifest disease have consistently yielded disappointing results^5^. Hypothetically, intervening in the prodromal phase of the disease may prove more effective^6^. A successful intervention could delay PD onset or -for some individuals-prevent progression to clinically manifest PD. There is broad consensus that disease modifying strategies are needed in the prodromal phase, with planning of the first trials now underway^7^.

### Nature of intervention

Exercise is a leading candidate for a scalable and feasible intervention, with converging evidence for potential disease-modifying effects in PD, alongside its well-established symptomatic benefits^11^. Specifically, aerobic exercise holds disease-modifying potential, as it attenuates motor symptoms^8-10^, potentially by promoting adaptive neuroplasticity in people with PD^11^, which aligns with preservation of basal ganglia networks as demonstrated in animal models^12^.

Because of the long duration of prodromal PD (up to 20 years or longer) and the lack of disease-specific progression markers, PD-prevention trials must be of sufficiently long duration to detect a clinically meaningful change. In a long duration study, maintaining compliance with increased levels of physical activity is important. Trials in clinically manifest PD, as well as other long-term prevention trials in other populations, have shown that exercise interventions are feasible for 6-24 months in trials that incorporated regular contact with study personnel^13,14^. While such an approach is feasible for studies with tens or hundreds of participants, regular in-person contact with a dedicated team is not a scalable approach for population-wide prevention. A promising novel approach is to replace in-person contact with digital engagement in lifestyle interventions delivered remotely using a motivational app (incorporating elements of ‘gamification’),^8,15^ and to also measure the outcomes remotely whenever possible, thus creating a fully ‘remote’ trial delivered in the participants’ own home environment. We explore this novel approach in the Slow-SPEED trials.

### Study populations

Several domains associated with prodromal PD have been identified^16,17^. Among the most well-established prodromal risk factors are isolated/idiopathic REM sleep behaviour disorder (iRBD), olfactory dysfunction (OD), and genetic predisposition (e.g., glucocerebrosidase 1 (*GBA1)* or Leucine-Rich Repeat Kinase 2 (*LRRK2)* variants). These groups carry a substantially elevated risk of PD, supporting the assumption that all groups represent prodromal PD and suitable target populations for prevention trials^18^. Further details regarding the prodromal risk groups can be found in Supplement 1

We outline the protocols for three trials which examine a shared remotely delivered physical activity intervention in complementary prodromal PD groups: individuals with iRBD (Slow-SPEED-NL, the Netherlands), those with OD (Slow-SPEED-UK, United Kingdom), and individuals with a genetic predisposition (Slow-SPEED-US, United States of America) (see Table 1).

**Table 1.** Comparison of the three Slow-SPEED trials. NL = the Netherlands, UK = the United Kingdom, US = the United States of America, iRBD = isolated REM sleep behaviour disorder (International Classification of Sleep Disorders, Third Edition - Text Revision), MRI = magnetic resonance imaging, Olfactory dysfunction = defined in this study as hyposmia corresponding to olfactory performance below the 15th percentile for age and sex on the University of Pennsylvania Smell Identification Test (UPSIT).

| **Trial** | **Slow-SPEED-NL** | **Slow-SPEED-UK** | **Slow-SPEED-US** |
| --- | --- | --- | --- |
| **Total participants** | 110 | 110 | 600 |
| **Duration** | 24 months | 18 months | 36 months |
| **Study population** | iRBD | Olfactory dysfunction | *LRRK2* G2019S or *GBA1* N370S carriers |
| **Intervention** | Physical activity | Physical activity | Physical activity |
| **Primary outcome(s)** | Step count | Step count | Step count & prodromal load |
| **Digital biomarkers** | Yes | Yes | Yes |
| **Blood biomarkers** | Yes | Yes (optional) | No |
| **Skin biopsy** | No | Yes (optional) | No |
| **Brain MRI** | Yes | No | No |

## Objectives

The Slow-SPEED trials evaluate the feasibility and preliminary efficacy of a long-term, remotely delivered, gamified physical activity intervention via the dedicated Slow-SPEED smartphone app.

The primary objective is to assess the feasibility of achieving a sustained increase in physical activity over 18 to 36 months through a remote motivational approach. The primary endpoint is change in daily step count from baseline to the last 12 weeks of planned follow-up. In Slow-SPEED-US, there is a co-primary objective examining the effect of increasing physical activity on the emergence of prodromal features of PD. This is a secondary objective in Slow-SPEED-NL and Slow-SPEED-UK.

The trials examine several other secondary objectives. These relate to:

- the feasibility of achieving and sustaining change in moderate-to-vigorous physical activity (MVPA);
- the feasibility of remotely monitoring digital biomarkers of motor- and non-motor features;
- the preliminary effect of physical activity on digital markers for physical fitness, motor- and non-motor features, and blood-, skin- and MRI-based biomarkers.

Figure 1 displays the logic model of the anticipated preliminary effect of increased levels of physical activity.

**Figure 1.**
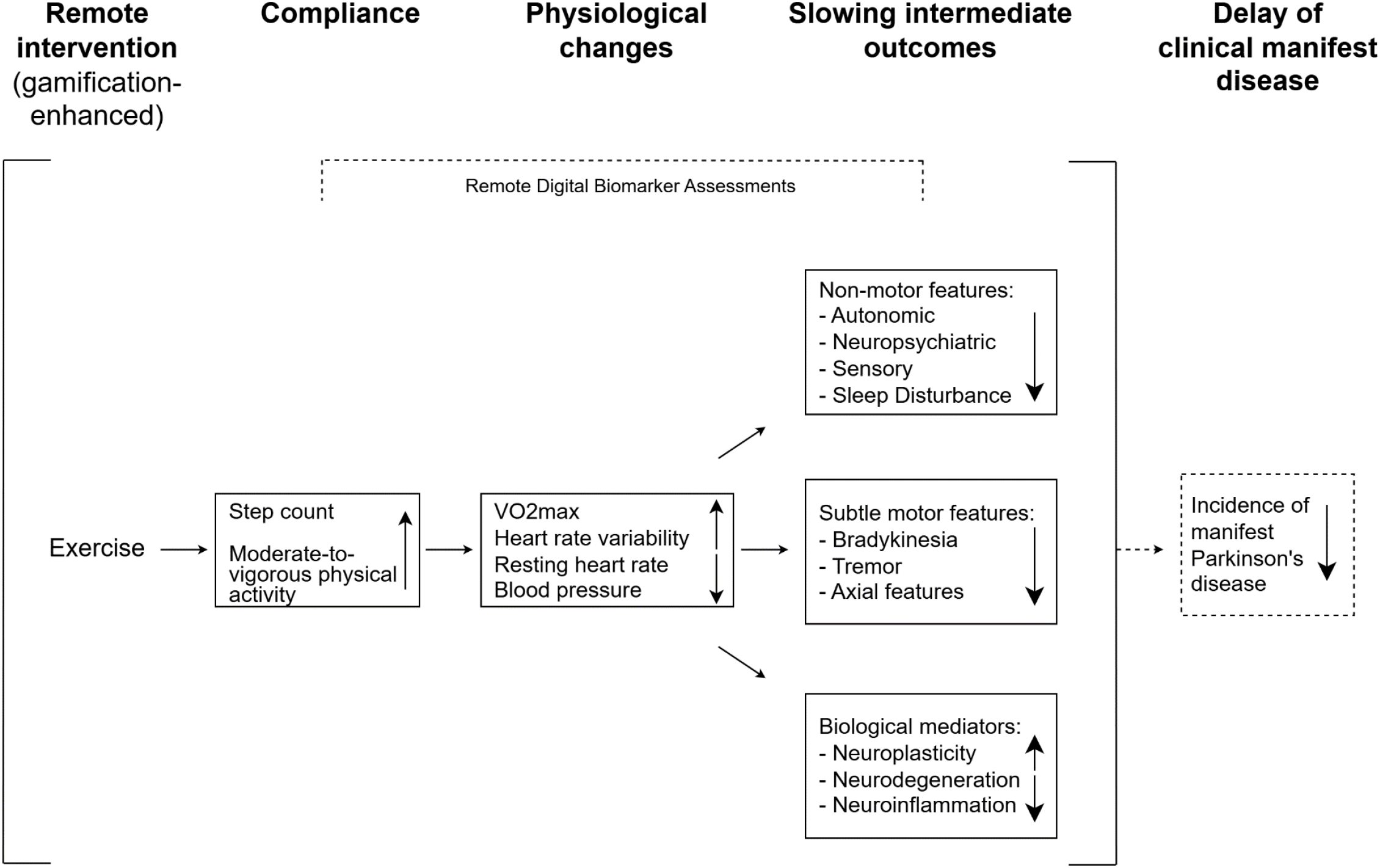
Logic model of all Slow-SPEED trials

### Trial design

The Slow-SPEED trials are an international effort comprising three double-blind randomised controlled trials in the Netherlands, the UK and the US that include an intervention and an active-control group. Figure 2 shows a schematic depiction of the intervention. Figure 3 shows the timing of outcome assessments for each trial. Although a longer follow-up of up to 36 months is desirable to assess longer-term effects, differences in available funding have resulted in varying trial durations. Follow-up time starts at randomisation and ends at the first of the following: end of planned follow-up, death, or loss to follow-up.

**Figure 2.**
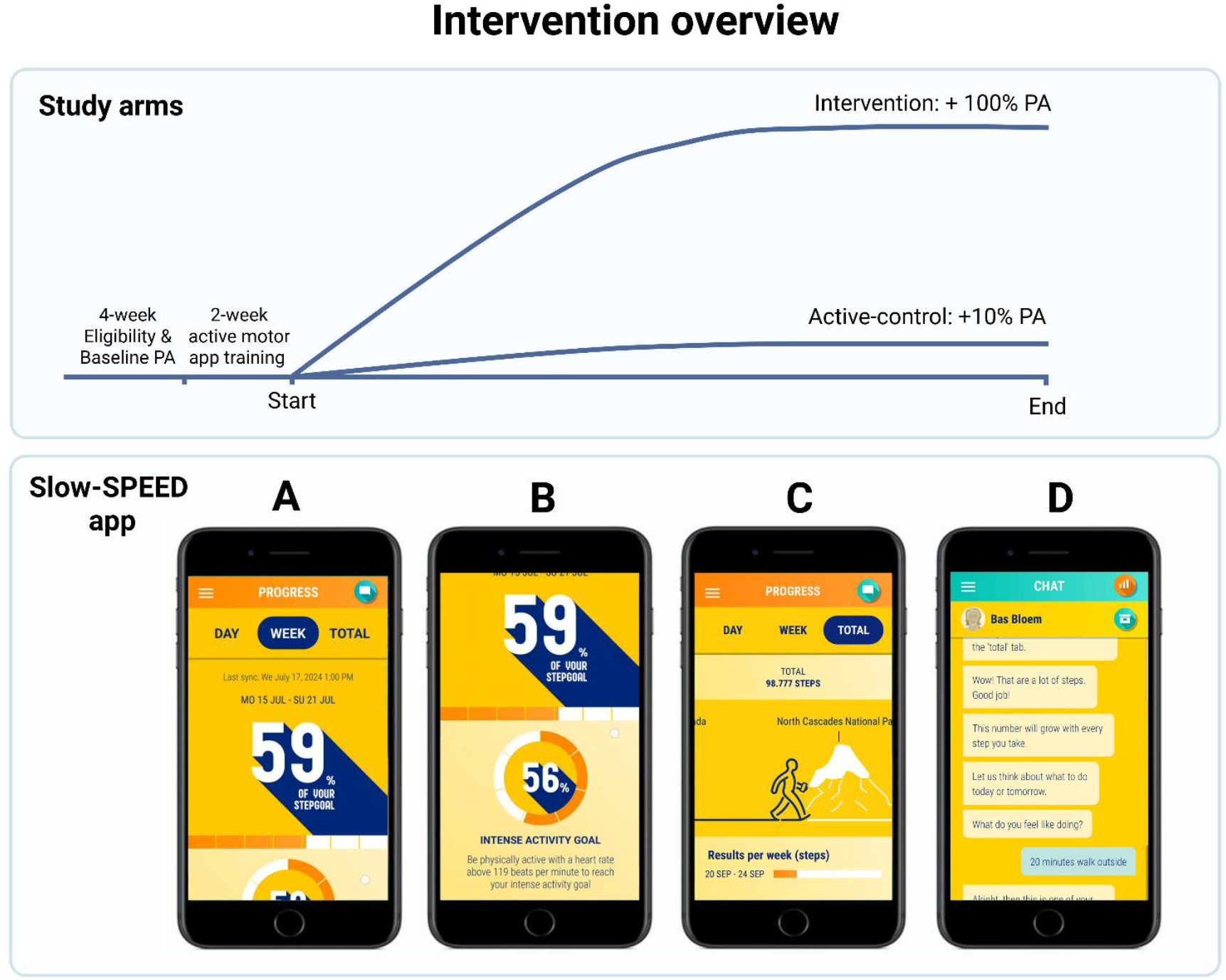
Intervention overview. Participants complete a screening by phone followed by a 4-week eligibility period to assess steps per day and minutes of moderate-to-vigorous physical activity (MVPA). If included, eligible participants attend the study centre for baseline assessments and receive instructions for home-based measurements (Slow-SPEED-NL & Slow-SPEED-UK), or receive instructions for home-assessments via a phone call without an in-person visit (Slow-SPEED-US). Following the baseline visit (Slow-SPEED-NL & Slow-SPEED-UK) or phone call (Slow-SPEED-US), additional home-based secondary outcomes are collected during a two-week period, which also includes a training phase with the Roche PD Research App to minimize potential learning effects. After this run-in phase, participants are randomised into the intervention or active-control arm. At the end of the study, participants in Slow-SPEED-NL and Slow-SPEED-UK return to the study centre for final in-clinic and home-based assessments, whereas participants in Slow-SPEED-US complete home-based assessments only, after which data collection is complete. PA = physical activity. Impression of the Slow-SPEED app. A: Progression towards personalised weekly step count targets; B: progression towards personalised weekly MVPA targets; C: total cumulative progression visualised as a virtual walking route through the United States or Europe, 15

**Figure 3.**
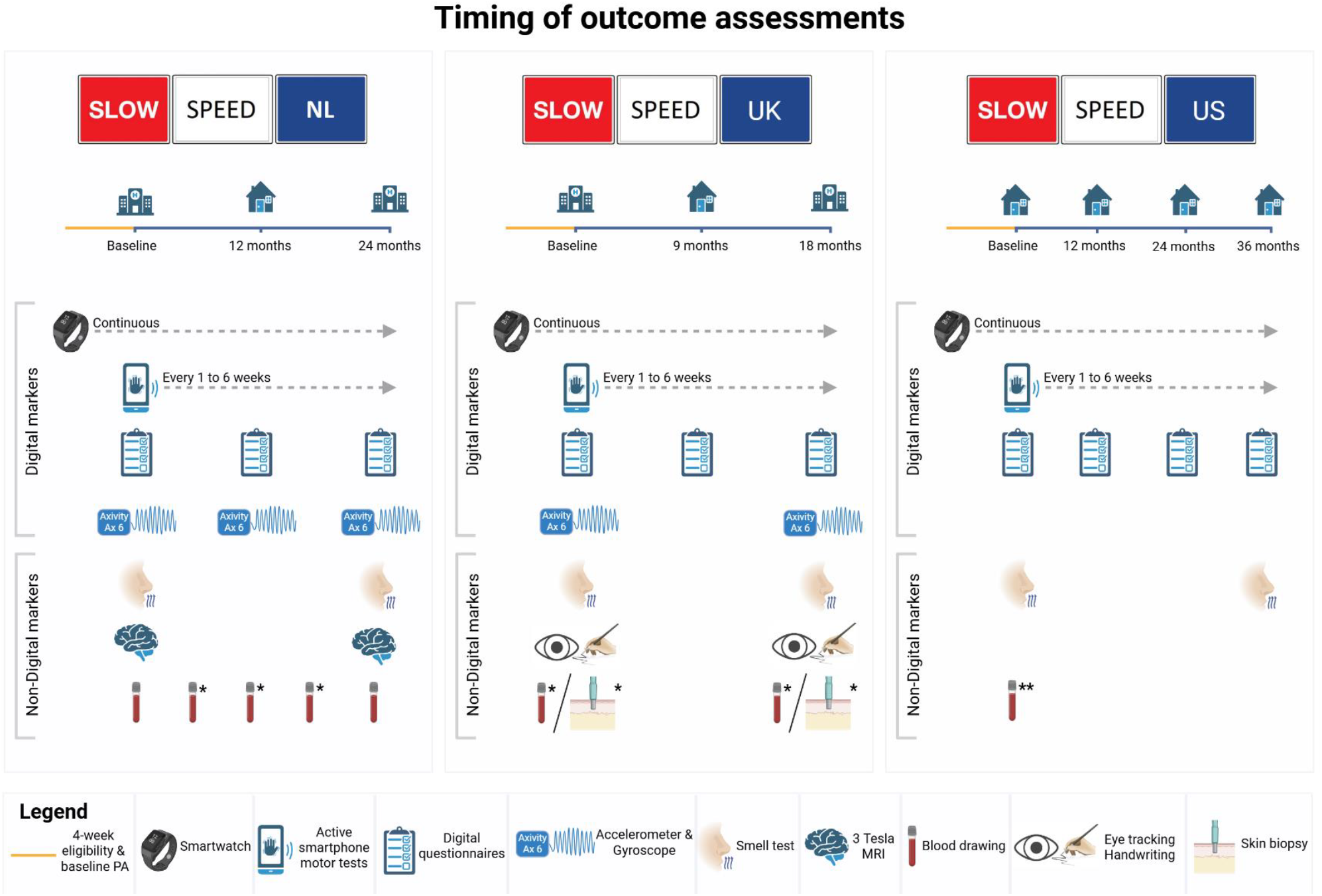
Timing of outcome assessments in the Slow-SPEED trials: From top to bottom: continuous monitoring with a wrist-worn activity tracker; motor assessments every 1–6 weeks using the Roche smartphone application; digital questionnaires at trial-specific time points; Periodic 7-day wrist-worn accelerometry and gyroscope assessments using Axivity AX6. In Slow-SPEED-NL, a lower-back Axivity AX6 is worn at baseline and end of study, with an additional assessment after 12 months in participants consenting to optional 6-monthly blood sampling. In Slow-SPEED-UK, wrist and lower-back Axivity AX6 are worn at baseline, 9-month follow-up, and end of study. In Slow-SPEED-UK, eye tracking movement and digital handwriting dynamics at baseline and end of study. Olfactory testing is performed at baseline and end of the study. In Slow-SPEED-NL, brain MRI scans are performed at baseline and end of study. Blood sampling are conducted at baseline and at the end of the study. *Indicates optional assessments. **Optional blood assessment may be used for future genetic analyses through the Global Parkinson’s Genetic Program (GP2).

## Methods: Participants, interventions and outcomes

### Study setting

This protocol was developed in accordance with the SPIRIT 2013 guidelines^19^. Slow-SPEED-NL is sponsored by and performed at Radboud University Medical Center (Radboudumc) and the Donders Center for Cognitive Neuroimaging (DCCN), Nijmegen. MRI-imaging is performed at DCCN. Slow-SPEED-UK is sponsored by and performed at Queen Mary University of London (QMUL). Slow-SPEED-US is sponsored by Radboudumc and is jointly coordinated by the University of Rochester and Radboudumc.

### Eligibility criteria and consent procedure

The eligibility criteria of the three Slow-SPEED trials differ regarding the group with a prodromal PD risk factor: Slow-SPEED-NL participants are required to have a video-polysomnography confirmed isolated RBD diagnosis as defined by the International Classification of Sleep Disorders, Third Edition, text revision (ICSD-3-TR)^20^; in Slow-SPEED-UK, participants are required to have hyposmia, defined based on performance on the 40-item British version of the University of Pennsylvania Smell Identification Test (UPSIT) (below the 15^th^ percentile for age and sex)^21^; in Slow-SPEED-US, participants are required to have a *LRRK2* G2019S variant or *GBA1* N370S variant, defined on the basis of prior genetic testing performed by 23andMe Research Institute, an institute that provides direct-to-consumer genetic analyses^22^. The interventional Slow-SPEED app is available in the Dutch and English language. In Slow-SPEED-NL the Dutch version is used, while in Slow-SPEED-UK and US trial the English version is used. The other inclusion criteria for each study are: aged 40 years or older (≥50 years for Slow-SPEED-US), able to walk independently, in possession of a suitable smartphone, average <7,000 steps/day (Slow-SPEED-UK or <10,000 steps/day during a 4-week eligibility period (Slow-SPEED-NL & Slow-SPEED-US).

The exclusion criteria of the three studies are identical: clinically diagnosed or self-reported diagnosis of manifest neurodegenerative disease; self-reported falls of three or more per year; dexterity or cognitive impairments hampering smartphone use; not community-dwelling; and unwillingness to receive risk information related to iRBD, OD or *LRRK2* or *GBA1* variant. Details of the eligibility criteria and exclusion criteria for the MRI assessments in Slow-SPEED-NL are provided in Supplement 2.

An overview and description of the consent procedure is shown in Figure 4. Before obtaining consent, we first evaluate PD risk awareness of interested participants^23^. This step is crucial to prevent potential harm that could arise from disclosing PD risk to those who are unaware. Further elaboration on the rationale for checking PD risk awareness can be found in Supplement 3.

**Figure 4.**
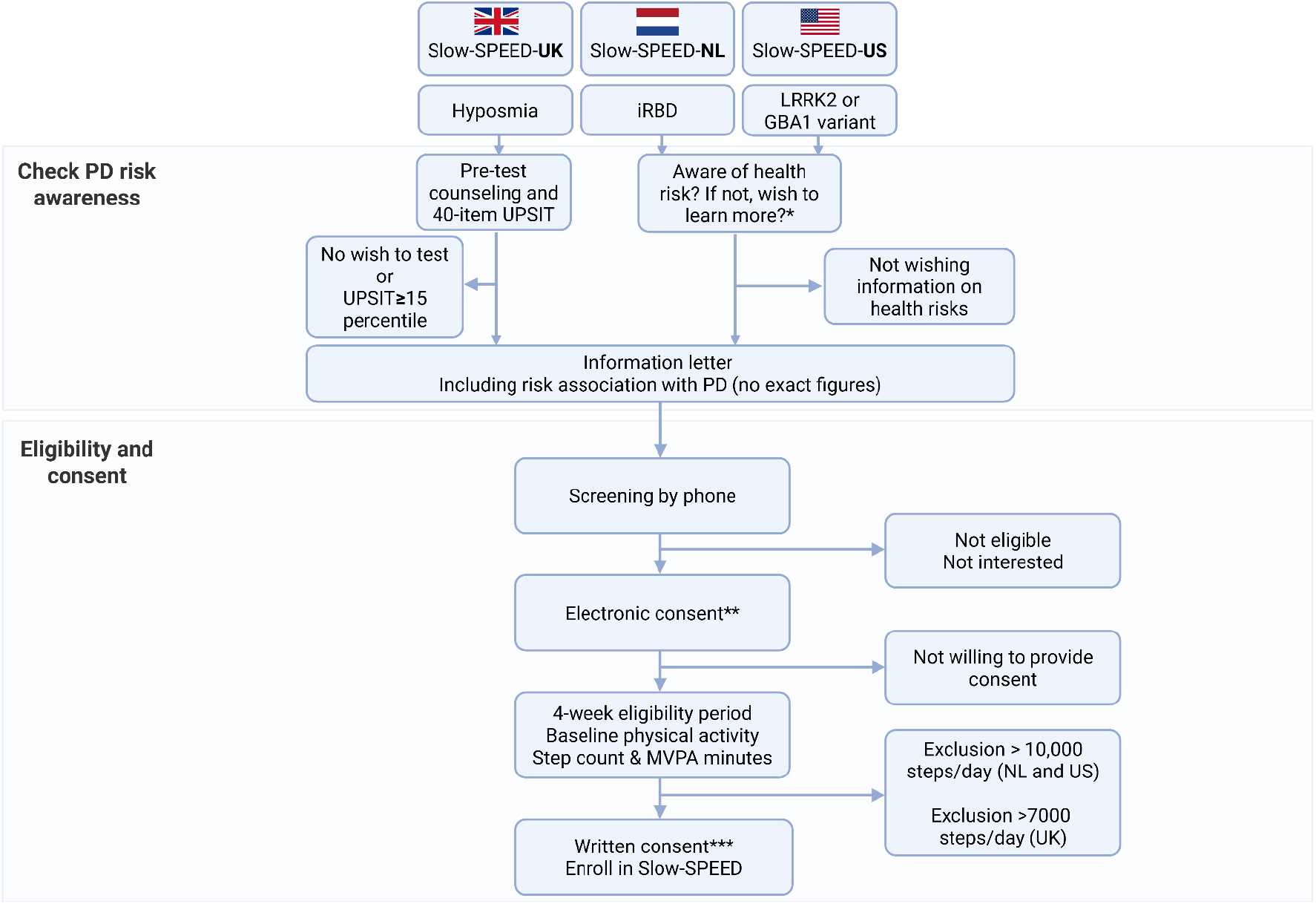
PD risk assessment and eligibility and consent procedure. After verifying that potential participants are aware of their PD risk or choose to learn about it through the study materials, they are screened by telephone for each trial’s eligibility criteria. Participants in Slow-SPEED-NL and Slow-SPEED-UK provide consent in a two-step procedure. First, they sign an electronic consent to allow data collection during the 4-week eligibility period. If they meet the activity-based inclusion criterion and can be enrolled in the study, they are asked to provide written informed consent at the start of their first study visit, before any study procedures take place. Informed consent is obtained by a trained researcher. We use this staged approach to minimise participant burden and respect their autonomy^43,44^. Participants in Slow-SPEED-US provide informed consent once electronically prior to the 4-week eligibility period, which also covers the entire study duration. All participants are provided with a copy of the written or electronically signed informed consent form. *NL participants are screened by phone; US participants are screened through a digital form **US participants provide electronic consent to cover both the 4-week eligibility period and intervention period. ***UK and NL participants provide electronic consent for the 4-week eligibility period followed by written consent at the baseline to cover the intervention period. PD = Parkinson’s disease; iRBD = isolated rapid eye movement sleep behaviour disorder; NL = Netherlands; UK = United Kingdom; US = United States of America; UPSIT = University of Pennsylvania Smell Identification Test; MVPA = moderate-to-vigorous physical activity.

## Interventions

### Intervention description

We refer to the intervention as ‘exercise’ for the structured, weekly nature of the energy-expending activities we encourage participants to engage in. For the outcomes, we use the term ‘physical activity’ as we aim to capture a broader range of behaviours, including both structured and unstructured physical activities throughout daily life.

The intervention comprises a smartphone app (Slow-SPEED app; Figure 2) that aims to motivate individuals at risk for PD to sustainably increase their physical activity levels. An earlier version of this same app was used successfully in persons with clinically manifest PD to enhance their levels of physical activity over the course of one year^15,24^. Primarily, the app motivates participants to take more steps and secondarily to engage in moderate-to-vigorous physical activity (MVPA). Walking is monitored through step count and MVPA is the time (in minutes) during which participants’ mean heart rate is ≥ 64% of their age-adjusted maximum heart rate, calculated as (HR_max_ = 207 – (0.7 * age))^25,26^. Both step count and MVPA are measured using a wrist-worn activity tracker^27^ (Fitbit model Charge 6, Google, Mountain View, CA, USA). Participants receive weekly masked targets for step count and MVPA. Both targets are measured independently.

The Slow-SPEED app incorporates various behavioural change techniques^28^ to help participants sustain increased physical activity levels (Supplement 4). Key features include receiving feedback on their weekly targets and support from a programmed virtual coach (without human interaction). The virtual coach sends messages, videos and audio recordings offering support, tips, and health education. By combining the step count and MVPA target visualization with the virtual coach, we aim to balance autonomy (allowing participants to decide when and how often they reach their targets) with the sense of remote supervision provided by the virtual coach^29^. Participants in the intervention group will ultimately be encouraged to increase from their baseline step count and MVPA levels -as determined in the 4-week eligibility period-by up to 100%. Those in the active control group are encouraged for an increase by up to 10%. As participants are excluded based on high baseline step counts, but not based on high baseline MVPA minutes, we set the MVPA week target to a maximum of 300 minutes per week for both study arms, twice the recommended number^25^. This approach prevents scenarios where participants with high baseline MVPA would face unrealistic targets. Participants in Slow-SPEED-NL have access to a Dutch version, while participants in Slow-SPEED-UK and Slow-SPEED-US have access to an English version of the app. Operationalization and details on the Slow-SPEED app are further described in Supplement 4.

## Outcomes

### Primary outcome in all three Slow-SPEED trials

The primary outcome in all three trials is the mean change in step count from the 4 weeks prior to baseline to the 12 weeks prior to the end of planned follow-up as measured with a wrist-worn activity tracker^27^. Step count was selected because it is an objective, continuously measurable, and clinically meaningful marker of physical activity that is sensitive to change, strongly associated with health outcomes, and feasible to capture remotely over long follow-up periods^30^. For each participant, we will calculate average daily step counts per four weeks throughout the study period.

### Additional co-primary outcome in Slow-SPEED-US

The co-primary outcome in Slow-SPEED-US is complemented by the mean change in an integrated *prodromal load score*, which summarizes digital motor and non-motor measures of PD, to quantify prodromal disease severity. The prodromal load score is currently being developed and will be described in a separate manuscript.

### Secondary outcomes

Secondary outcomes and their assessment schedule for each trial are presented in Table 2. A key secondary endpoint is MVPA, defined as minutes during which mean heart rate is ≥64% of age-adjusted HRmax, as recorded by the wrist-worn activity tracker. Other secondary outcomes include blood biomarkers of metabolism, inflammation, growth factors, aging, pathological proteins and neurodegeneration (Slow-SPEED-NL & Slow-SPEED-UK); optional skin-punch biopsies (Slow-SPEED-UK only); brain MRI measures (Slow-SPEED-NL only); and active and passive digital measures of motor functioning (all trials). Active monitoring is performed using the Roche PD Research app, a smartphone application that captures PD measures through task-based assessments, including gait, tremor, dexterity, and bradykinesia^31^. Details on the secondary outcomes are listed in Supplement 5.

**Table 2.**
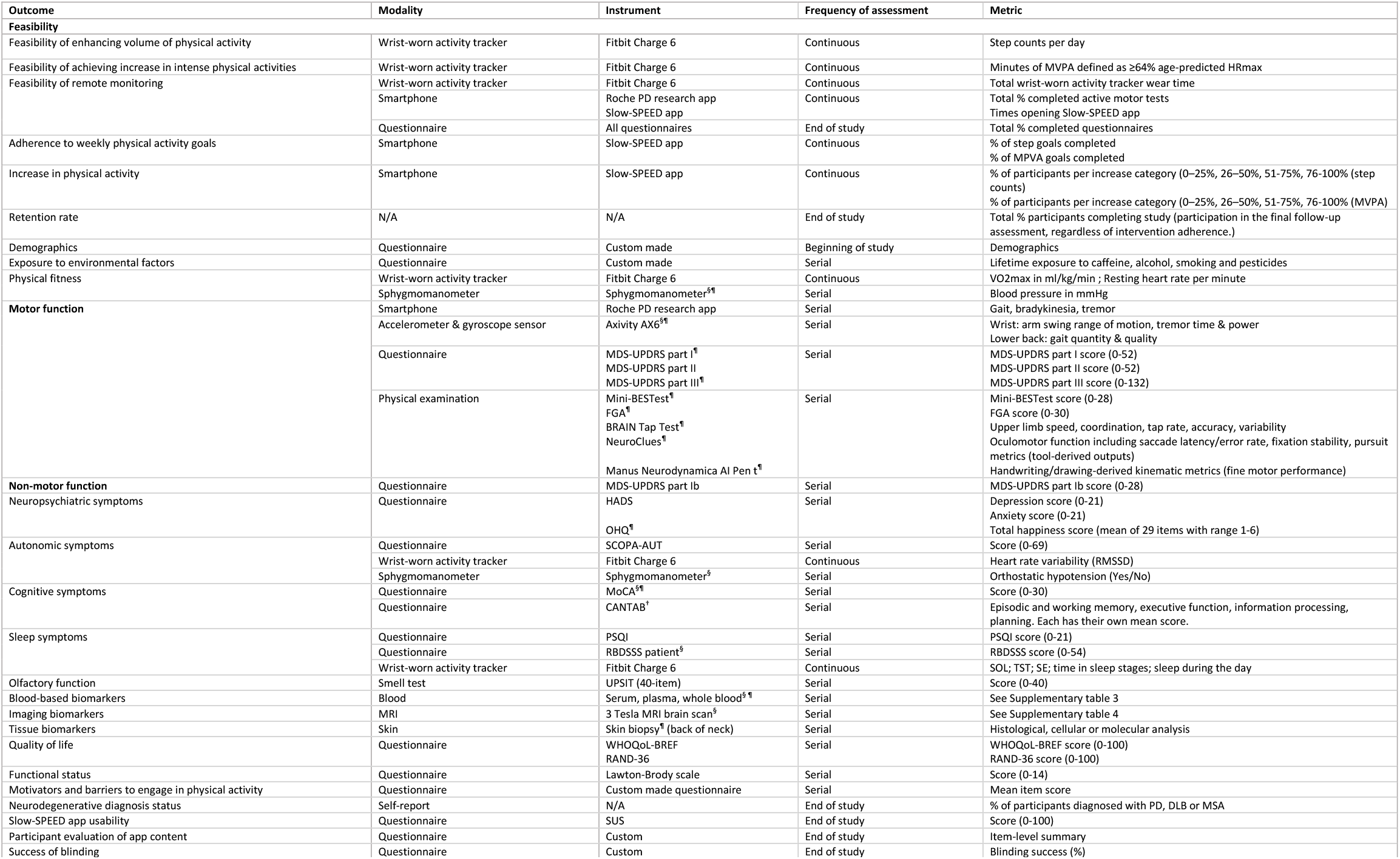
Overview of outcome assessments methods. All assessments are conducted in all three trials, except where noted: § = Slow-SPEED-NL only; ¶ = Slow-SPEED-UK only; † = Slow-SPEED-US only. BRAIN = BRadykinesia Akinesia Incoordination Tap Test; CANTAB = Cambridge Neuropsychological Test Automated Battery; DLB = dementia with Lewy bodies; FGA = Functional Gait Assessment; HADS = Hospital Anxiety and Depression Scale; MDS-UPDRS = Movement Disorders Society Unified Parkinson’s Disease Rating Scale; Mini-BESTest = Mini Balance Evaluation Systems Test; MoCA = Montreal Cognitive Assessment; MRI = magnetic resonance imaging; MSA = multiple system atrophy; MVPA = moderate-to-vigorous physical activity; N/A = not applicable; PSQI = Pittsburgh Sleep Quality Index; RAND-36 = RAND 36-Item Health Survey; RBDSSS = REM-sleep Behavior Disorder Symptom Severity Scale; RMSSD = root mean square of successive differences; SCOPA-AUT = Scales for Outcomes in Parkinson’s Disease - Autonomic Dysfunction; SOL = sleep onset latency; SUS = System Usability Scale; TST = total sleep time; SE = sleep efficiency; UPSIT = University of Pennsylvania Smell Identification Test; VO2max = maximum oxygen consumption rate; WHO-QoL-BREF = World Health Organization Quality of Life Instrument - Brief.

### Participant timeline

After confirming PD-risk awareness and screening by phone, participants begin a four-week eligibility screening period during which baseline step count and minutes of MVPA per day are determined, followed by baseline assessments and randomisation into the intervention or active-control arm (Figure 2). See Figure 3 for an overview of the timeline of assessments for each trial.

### Sample size and recruitment strategies

The intended total sample size of each of Slow-SPEED-NL and Slow-SPEED-UK is 110 participants based on the primary endpoint of change in step count. The intended sample size of Slow-SPEED-US is 600 participants based on the co-primary endpoints of change in step count and change in prodromal load score. See Supplement 6 for further elaboration on the sample size calculation and recruitment strategies.

## Treatment assignment, blinding and participant retention

Further elaboration on treatment assignment, blinding, criteria for discontinuing or modifying allocated interventions; strategies to improve adherence to interventions and promote retention, and relevant concomitant care permitted or prohibited during the trial are provided in Supplement 7.

## Data collection and management

Elaboration on data management and confidentiality can be found in Supplement 9.

## Statistical methods

Data will be analysed according to the intention-to-treat principle. Sensitivity analyses will include a per protocol analysis.

### Primary outcome

The primary study outcome in each trial is the within-subject change in step count from the four weeks prior to baseline to the 12 weeks prior to the end of follow-up. To address the feasibility of sustainably increasing physical activity using a remote physical activity intervention, we will compare the intervention and the active control group. We will calculate average daily step counts for each participant every four weeks throughout the entire study period, which includes the four-week period prior to baseline and the 104-week treatment period in Slow-SPEED-NL (total 27 time points), 78-week treatment period in Slow-SPEED-UK (total 20 time points), and 156-week treatment period in Slow-SPEED-US (total 40 time points).

We will use a mixed-effects model in which the dependent variable is the average daily step count in each four-week interval. The model will be implemented as a constrained longitudinal data analysis in which average baseline step counts are equal across treatment groups^32^. This reflects the sampling of all participants from a single population prior to randomization and has the statistical benefit of permitting inclusion of all participants regardless of missing data and of removing variance attributable to each participant’s baseline step count in a manner similar to analysis of covariance^33^.

The primary analysis will not assume a specific pattern of change in average step count over time. The model will include fixed effect terms for time (categorical by four-week interval), treatment group * pre/post treatment initiation * time, and age, sex and their interaction with time. In Slow-SPEED-UK, the fixed effects will also include terms for randomisation strata (age group, baseline step-count group, and UPSIT-defined hyposmia severity) and their interactions with time. To account for within-person correlations in step counts over time, the model will include participant-level unstructured covariance. Simpler covariance structures, guided by data from the STEPWISE trial, will be used if the model with unstructured covariance fails to converge^24^.

The primary contrast will estimate the treatment-dependent difference in step counts over the final 12 weeks of follow-up compared with the four weeks prior to baseline using least-square means. A positive, statistically significant increase in step count in the intervention group relative to the active-control group would indicate a sustained increase in physical activity.

Subgroup analyses will be conducted stratified by baseline daily step counts (three levels: <5,000, 5,000-6,999, and 7,000-9,999 steps per day) by including additional subgroup, subgroup * time, and subgroup * treatment group * pre/post treatment initiation * time fixed effect terms. Analyses will be repeated after pooling data across the three trials and including trial, trial * time, and trial * treatment group * pre/post treatment initiation * time fixed effect terms with estimates at the common 18-month time point averaged across trials.

### Joint primary outcome US trial

The co-primary outcome in Slow-SPEED-US is the within-subject change in prodromal load score. This will be analysed using the same mixed-effects model described above for step count.

### Secondary outcomes and interim analyses

The statistical analysis of the secondary outcomes and pre-specified interim analyses for each trial are described in Supplement 10.

### Oversight and monitoring

Information regarding the composition of the coordinating centres and trial steering committee, adverse event reporting and harms and plans for communicating important protocol amendments to relevant parties is described in Supplement 8.

## Discussion

We describe the first long-term non-pharmacological trials conducted in populations suspected to be in the prodromal phase of PD. These trials will provide valuable evidence on the feasibility and preliminary efficacy of a remote, smartphone-based physical activity intervention for PD prevention.

Trials focused on the prevention of PD are rare, with only one trial currently active^34^. Two additional pharmacological trials^35,36^, as well as the Path To Prevention (P2P) platform trial nested within the Parkinson’s Progression Markers Initiative (PPMI), are currently in preparation^37^. The Slow-SPEED studies have several strengths. First, the studies will provide insights into the feasibility of remotely delivering an intervention and remote monitoring of outcomes in at-risk individuals. Such a remote trial design is likely to enhance compliance, which is particularly needed for the typically long-term trials that are needed to demonstrate disease modification in the prodromal phase of PD. Additionally, this remote approach is potentially scalable to a range of settings, including to low-resource settings. The knowledge that will be gained from this approach could serve as a blueprint for future PD prevention trials, including trials of pharmacological or dietary interventions. Second, these are the first trials to examine a non-pharmacological intervention in relation to the possible prevention of PD. Previous attempts to delay clinically manifest PD using pharmacological interventions have been unsuccessful, whereas exercise has shown greater potential based on both animal work, observational epidemiological studies and clinical trials. Compared to studies of drugs (which typically have a monomorphic mode of action), nonpharmacological interventions such as exercise may potentially hold greater promise of being able to successfully delay neurodegeneration because of their more pleiotropic and multifaceted mode of action, which simultaneously tackles multiple different elements of the complex underlying pathophysiology of PD^38,39^. As such, these studies will offer unique insights into the preliminary efficacy of physical activity to delay progression of digital measures and fluid- and neuroimaging biomarkers of prodromal PD. Third, the study outcomes are monitored over 18 to 36 months, including several outcomes which are continuously monitored, providing long-term insights into both the natural course and potential effects of physical activity over long intervals. Fourth, the trials will examine complementary prodromal populations, including individuals with iRBD, OD, and those at genetic risk for PD. Fifth, the harmonised design (shared devices and endpoints) across three countries enables pre-specified individual-participant-data meta-analyses and cross-cohort comparisons, which may improve precision and generalisability. Sixth, the Slow-SPEED trials establish reusable methodologies for decentralised prevention trials (e-consent, remote monitoring, digital coaching), positioning the program as a platform for larger, future studies.

Apart from the strengths, we also anticipate some challenges. First, we will determine whether it is feasible and ethically responsible to include participants at risk for PD remotely. To date, no such trials have been completed. However, we anticipate that inclusion will be feasible thanks to strong collaborations with sleep expertise centres, the ongoing observational study PREDICT-PD and the 23andMe Research Institute. 23andMe currently has a participant base of more than 40,000 non-manifest carriers of a *LRRK2* G2019S or *GBA1* N370S variant. Additionally, by using tailored strategies for risk disclosure and informed consent, we believe that we can protect participants from adverse psychosocial effects of learning potentially unwanted information about future disease risks.

Furthermore, as the majority of our source populations are drawn from existing research cohorts and participation requires voluntary enrolment, we anticipate underrepresentation of individuals from minority ethnic backgrounds and those with lower socioeconomic status or health literacy.

Second, participants may not adhere to their assigned interventions, as lifestyle changes are known to be intrinsically hard to sustain^40^. The main challenge for participants will be to stay more physically active over a fairly long time span supported solely by a remote intervention that offers little to no human interaction. Yet, we believe that long-term adherence is achievable, thanks to the diverse behaviour change techniques integrated into the app. This confidence is grounded in our extensive experience of high compliance with a similar app used in the STEPWISE trial involving individuals with clinically manifest PD^15,24^. To mitigate decline in adherence, we pre-specify engagement supports (e.g., regular updates, virtual events, gym memberships and maps through Ordnance Survey app for outdoor walking routes (in the UK), and reimbursement of travel costs where applicable. Also, although evidence suggest the greatest benefit for PD symptoms from vigorous exercise, we selected a moderate-to-vigorous intensity cut-off for its practicality and broad applicability, particularly given the remote nature of the trial and the slightly increased health risks when focusing on vigorous exercise only^41,42^. Third, we use age-predicted maximal heart rate to define each participant’s MVPA heart-rate cut-off point. This approach does not account for rate-limiting/chronotropic medications or autonomic dysfunction.

Fourth, the Slow-SPEED trials do not incorporate dopaminergic-imaging or assessment of biomarkers in cerebrospinal fluid primarily due to cost and logistical constraints. Nonetheless, we believe that the fluid and imaging biomarkers included in a subset of participants will help to characterise potential biomarkers of prodromal PD and the effects of exercise on them. Fifth, we do not apply additional prodromal criteria beyond the primary risk factor for each trial. Consequently, a subset of participants may not develop prodromal PD throughout the trial, limiting our ability to detect progression. This consideration may be particularly relevant for non-manifest gene variant carriers.

Sixth, potential sources of bias may be present (e.g., seasonal variation in activity, not wearing digital devices, contamination via outside exercise programmes). To mitigate bias, these will be addressed with robust data-quality rules and mixed-effects models that adjust for baseline activity and time.

These trials are the first large-scale, long-term, remote, randomised controlled trials to investigate the feasibility of increasing physical activity in individuals in the prodromal phase of PD and preliminary efficacy of physical activity for prodromal disease progression. The shared data dictionary facilitates pooled analyses across sites, accelerating learning and de-risking the future design of subsequent adequately powered prevention trials. Together, these features position Slow-SPEED to evolve into a platform that can test multiple interventions and at-risk populations over time. In summary, the inclusion of three distinct prodromal PD populations across multiple countries, combined with continuous remote monitoring and the incorporation of neuroimaging, skin punch and blood-based biomarkers, demonstrates the feasibility of a scalable approach for future PD prevention studies aimed at modifying prodromal disease progression. Building on this work, the Slow-SPEED programme may be extended to additional settings in the coming years.

## Trial status, data availability and dissemination

The full protocols of the Slow-SPEED trials are registered at ClinicalTrials.gov (NCT06193252; NCT06600438; NCT06993142). The first Slow-SPEED-NL participant was randomized on June 7th, 2024. We expect to randomize the last participant in the trial in the Netherlands by the end of 2026. As of December 19^th^, 2025, the latest version of the protocol was version 8.0 (January 15^th^, 2025). The Slow-SPEED-UK protocol is currently under review by an Institutional Review Board to obtain ethical approval (IRAS ID 362207). The first Slow-SPEED-US participant was randomized on November 24^th^, 2025. As of December 19^th^, 2025, the latest version of the protocol was version 1.0 (April 17^th^, 2025).

Publications arising from these trials will be available via Open Access where possible. Data generated from these trials will be shared in an online repository for collaborative sharing after the main results of each trial have been published, and upon application to, and approval by, the trial management group. Applicable agreements will be signed by the involved parties. Findings will be disseminated through peer-reviewed publications, scientific conferences, and plain-language summaries shared with participants and relevant patient and public organisations.

## Data Availability

This manuscript concerns a clinical trial design protocol. No data has currently been produced.

## Acknowledgements

We like to thank the research participants and employees of sleep centres Kempenhaeghe and Stichting Epilepsie Instellingen Nederland (SEIN), PREDICT-PD and 23andMe Research Institute. We are especially grateful to Paul Cannon for his valuable input in shaping recruitment strategies in collaboration with 23andMe Research Institute. We acknowledge the contributions of the research coordinators, data managers, and clinical staff at all participating sites for their invaluable support in recruitment and data collection. Finally, we appreciate the input from patient representatives, whose perspectives helped shape the design and feasibility aspects of this trial.

We gratefully acknowledge the invaluable contributions of individuals with lived experience of prodromal or early-stage Parkinson’s disease, as well as members of the public, who supported the development of the three Slow-SPEED trials. Their insights informed the study design, participant-facing materials, and overall trial delivery. We especially thank those who provided feedback on recruitment materials, informed consent procedures, and communication approaches, helping us ensure non-alarmist framing of Parkinson’s risk and minimise participant burden. Their input directly shaped several key design decisions, including the fully remote and unsupervised study format, the adoption of gradual and individualised physical-activity targets, and the usability and motivational features of the gamified smartphone application. We also thank all participants who engaged with ongoing trial communications and offered feedback on procedures, acceptability, and user experience. Their continued involvement has been essential in refining and improving the conduct of the trials.

These studies are funded by ZonMW (Veni; 09150162010183), ParkinsonNL (P2022-07), Parkinson’s Foundation (PF-Trail-1421986), Michael J Fox Foundation (MJFF-022767), the Dutch Research Council (ERA4HEALTHNUTRIBRAIN-89), Davis Phinney Foundation (no grant number), Edmond J Safra Foundation (no grant number), Donders Institute (in kind contribution), Parkinson’s UK (Grant Award J-2301), Cure Parkinson’s (no grant number). The funders had no role in the study design, data collection or in the writing of the manuscript. Any material involvement of a funder beyond financial support would be explicitly declared. None occurred.

## Author contributions

All authors contributed to writing the manuscript. THO, EM and VA wrote the first draft of the manuscript. THO, EM, VA, BRB, RBS, AJN and SKLD designed the studies. BRB, RBS, AJN and SKLD supervise the projects and provide guidance. All authors contributed substantially to the conception of the studies, take full responsibility for the accuracy and integrity of the work, and approved the final version for submission.

