## Supplementary materials for "Slow-SPEED: protocol for three randomised trials of remotely delivered exercise to prevent Parkinson’s disease"

### Supplement 1 – Justification of the study populations

**Isolated/idiopathic REM sleep behaviour disorder (iRBD)**
Video-polysomnography confirmed iRBD is the strongest predictor of clinically manifest disease, with an estimated annual conversion rate of 6.3% and greater than 70% risk of progressing to PD or a related neurodegenerative disease within 12 years[1]. Individuals with iRBD are of high interest for recruitment in upcoming PD prevention trials. However, an estimated two-thirds of individuals with early manifest PD do not present with iRBD, underscoring the importance of studying complementary prodromal groups as well[2].

**Hyposmia**
To diversify our study population, we also include individuals with olfactory dysfunction (OD), the most common prodromal feature associated with PD. In Slow-SPEED-UK, OD defined as hyposmia based on UPSIT performance. Hyposmia is associated with approximately a 4-fold (95%CI [2.47-7.07]) increased risk of developing PD[3]. However, hyposmia is nonspecific, as the majority of hyposmic individuals in community cohorts do not develop PD or related synucleinopathies[4]. This highlights the need to combine hyposmia with additional biomarkers to enrich for individuals at increased risk.

**Genetic predisposition**
In addition to prodromal symptoms, several genetic groups are being considered as target populations for prevention trials of PD. We focus on two specific strata, namely carriers of the *LRRK2* G2019S variant – which has a penetrance of around 30% by age 80 but is rare in the general population– and carriers of the *GBA1* N370S (also known as N409S) genetic variant, which has a penetrance of less than 10% but is much more common[5-8]. A longitudinal study of individuals who carry these variants can add valuable insights for understanding different prodromal disease trajectories. Furthermore, it is important to include individuals with a genetic predisposition, as observational studies indicate that the association between physical activity and PD risk may differ based on genotype[9].

#### Supplement 2 – Eligibility criteria Slow-SPEED trials

| **Eligibility criteria** | **Slow-SPEED-NL** | **Slow-SPEED- UK** | **Slow-SPEED-US** |
|---|---|---|---|
| **Inclusion criteria** | | | |
| 50 years or older | | | X |
| 40 years or older | X | X | |
| Understands Dutch language | X | | |
| Understands English language | | X | X |
| Able to walk independently inside the home without the use of a walking aid. | X | X | X |
| In possession of a suitable smartphone for Slow-SPEED, Fitbit and Roche PD research app | X | X | X |
| Less than an average 10,000 steps/day during the 4-week eligibility period | X | | X |
| Less than an average 7,000 steps/day during the 4-week eligibility period | | X | |
| iRBD meeting ICSD-3-TR criteria[10] | X | | |
| Hyposmia (UPSIT <15$^{th}$ percentile for age and sex)[11] | | X | |
| *LRRK2* G2019S or *GBA1* N370S variant | | | X |
| **Exclusion criteria** | | | |
| Clinically diagnosed or self-reported diagnosis neurodegenerative disease | X | X | X |
| Self-reported falls 3 or more per year | X | X | X |
| Dexterity problems or cognitive impairments hampering smartphone use | X | X | X |
| Wish not to be informed about an increased risk of developing diseases associated with iRBD, hyposmia or LRRK2 or GBA1 variant | X | X | X |
| Not community-dwelling | X | X | X |
| Smartphone is one the following devices: Huawei P8 Lite ; Huawei P9 Lite ; Xiaomi Mi 6 ; Huawei P20 Lite (not compatible with the Fitbit platform) | X | X | X |
| **MRI exclusion criteria** | | | |
| History of epilepsy, structural brain abnormalities (e.g., stroke, traumatic defects, large archnoid cysts) or brain surgery | X | | |
| Claustrophobia | X | | |
| Implanted electrical device (e.g., pacemaker, DBS, neurostimulator) | X | | |
| Metal implant (e.g., prosthetics, ossicle prosthesis, metal plates, other non-removable metal part) | X | | |
| Pregnancy | X | | |

**Supplementary Table 1.** Eligibility criteria for all three Slow-SPEED studies. DBS = Deep brain stimulation; ICSD-3-TR = International Classification of Sleep Disorders Third Edition-Text Revision; iRBD = idiopathic REM-sleep behaviour disorder; MRI = Magnetic Resonance Imaging; UPSIT = University of Pennsylvania Smell Identification Test.

##### Rationale for age and step count differences between trials

In Slow-SPEED-NL, we initially included participants aged 50 years and older. To improve recruitment feasibility and enhance inclusivity, we later lowered the minimum age to 40 years. Based on this experience, Slow-SPEED-UK starts with recruiting participants aged 40 years and older. In contrast, Slow-SPEED-US maintains the threshold at 50 years and older, reflecting differences in population characteristics—specifically, a lower genetic risk for PD and a reduced likelihood of prodromal features emerging during the trial period as age decreases, compared to the risk strata used in Slow-SPEED-NL and Slow-SPEED-UK.

For eligibility, participants in Slow-SPEED-NL and Slow-SPEED-US must average <10,000 steps/day during the 4-week eligibility period. An initial cut-off of <7,000 steps/day was chosen to reduce ceiling effects, enhance sensitivity to detect intervention-related changes, and to ensure recruitment of a truly low-active population whom are most likely to benefit[12]. However, during early recruitment for Slow-SPEED-NL, we observed that most participants exceeded 7,000 steps per day at baseline. Consequently, the inclusion criterion was revised from <7,000 steps/day to <10,000 steps/day to maximize feasibility and inclusivity across a broad spectrum of activity levels[13]. In contrast, Slow-SPEED-UK initially retains the stricter inclusion criterion of <7,000 steps/day, but this may be broadened to <10,000 steps/day if participant activity levels mirror those observed in the Netherlands.

#### Supplement 3 – PD risk awareness and disclosure

For potential participants, receiving information about an increased risk of developing Parkinson's disease (PD) can be distressing and impactful, and if they are currently unaware of any risk of developing PD, they have a right not to know[14]. In the Netherlands, individuals with iRBD will be recruited from sleep centers. In the UK, participants with olfactory dysfunction will be recruited from the existing PREDICT-PD cohort via specialist Ear, Nose and Throat clinics[15]. In the US, *LRRK2* and *GBA1* carriers will be recruited via the 23andMe Research Institute that provides direct-to-consumer genetic testing and opportunities for members to participate in research studies.

To prevent unintentional risk disclosure, tailored strategies – modelled on existing guidance for the disclosure of PD risk[16]– will be used to confirm, before enrolment, that potential participants are already aware of their increased PD risk.

***Strategy for potential participants with iRBD***

Individuals with iRBD are routinely informed about their risk of PD by their neurologist. They are typically told the relationship between iRBD and PD in general, non-quantitative terms. Prospective participants who – when they are approached about the study by their sleep center – express an interest in Slow-SPEED-NL, will be informed about the study by telephone. During this phone call, a trained researcher will assess the participant's awareness of the health risks associated with iRBD. The researchers are trained by the study doctor, who is a neurologist with extensive experience with the target population. If participants are already aware of their future disease risks, the trained researcher will proceed with providing information about the study. If participants are not aware, the trained researcher will ask whether they wish to learn about the associated health risk. Those who agree will receive general, non-quantitative information about the increased risk of PD associated with iRBD, and may proceed to enrolment. Those who decline to learn more will be excluded from the study, to avoid potential harm from disclosing unwanted information about future disease risks.

***Strategy for potential participants with olfactory dysfunction from PREDICT-PD and Ear, Nose and Throat clinics***

Potential participants with olfactory dysfunction (OD) will be identified through the PREDICT-PD platform using an abbreviated 6-item smell test, and individuals identified through participating Ear, Nose and Throat Smell & Taste clinics, are not routinely informed of any increased risk of PD as part of standard clinical care or cohort procedures. OD is common (1in 5) and non-specific, and most individuals with smell loss will not develop PD. Individuals who express interest in Slow-SPEED-UK are invited to an in-person screening and consent visit. The study is presented as a feasibility trial of a remote physical-activity intervention in people with reduced sense of smell. Participants are asked, using an opt-in approach, whether they wish to receive general, non-quantitative information about possible health associations of OD. If requested, this information is provided in broad, non-alarming terms and emphasises uncertainty and heterogeneity. Participants who decline may continue screening without disadvantage. Following informed consent, eligibility is confirmed using the 40-item University of Pennsylvania Smell Identification Test (UPSIT). Hyposmia, defined by UPSIT performance below the 15th percentile for age and sex, is used solely for study eligibility and is not framed as diagnostic or predictive of PD. Participants meeting the eligibility criteria will proceed to enrolment.

***Strategy for potential participants with genetic variants***

Users of genetic testing of the 23andMe Research Institute routinely received PD risk information as part of their genetic health risk report. To users who carry variants in the *LRRK2* or *GBA1* genes, information is made available that indicates that 25% of individuals with *LRRK2* gene will develop PD. Information when carrying a *GBA1* gene variant include that there is a slightly increased risk, where the exact estimate is not available. Participants from 23andMe will only be contacted if 23andMe confirms that they have previously reviewed their PD genetic test results. Slow-SPEED-US, risk awareness screening is completed via a digital registration form. This includes a question to assess the participant's awareness of the health risks associated with the gene variant they carry. If participants are aware of their future disease risks, they will be able to continue registration to receive the study information form. If they are not aware of future disease risks, then participants will be notified that they may encounter information about these health risks during the study. A follow-up question includes asking whether they wish to learn these risks during the study. Those who agree will be able to continue registration. Those who decline to learn more will be excluded from the study, to avoid potential harm from disclosing unwanted information about future disease risks.

#### Support comprehension of PD risk prior to study enrolment

In all three trials, eligible participants will receive study information via email, explicitly stating that one of the study's aims is to monitor outcomes related to PD. This ensures that participants are aware that they may be confronted with individual research results related to PD during the study. In all trials, participants are informed that the study does not provide personalised, quantitative, diagnostic, or predictive information about PD risk. To monitor the wellbeing of participants, a follow-up phone call will be offered, in which the researcher checks whether participants have adequate understanding of their increased risk of PD and have not been harmed by this information.

This call focuses on confirming understanding that their risk factor for inclusion OD or the genetic variant is relatively common and non-specific, reinforcing the right not to receive further PD-related information, and identifying any distress arising from study discussions. If needed, participants may be referred to psychosocial support services. The same process is followed for individuals with iRBD, acknowledging that iRBD carries a higher risk of progression to PD than OD or genetic risk factors. This phone call also serves to support participant understanding of the study and to answer any remaining study-related questions.

#### Supplement 4 – Details on the Slow-SPEED app

##### Behavioural change techniques and virtual coach

Several behavioural change techniques (BCT) are implemented in the Slow-SPEED app (supplementary table 2 according to the BCT taxonomy v1[17]). The virtual coach is an automated chat function without direct human interaction. Participants can reply to the messages from the virtual coach with pre-set answer options, which prompts tailored replies from the coach based on their answer provided. Messages are initially sent every three days for the first 70 days, then shift to every six and twelve days for the remainder of the trial. All content is carefully designed for each country's population.

| BCT code and name | App component | Details |
|---|---|---|
| **1.Goals and planning** | | |
| 1.1 Goal setting (behaviour) | Progress screen | Weekly step count and MVPA targets are set and directly tied to the desired behaviour |
| 1.4 Action planning | Progress screen | The weekly targets include the duration and intensity of the desired behaviour |
| **2. Feedback and monitoring** | | |
| 2.2 Feedback on behaviour | Virtual coach | Adjusts messages based on whether participants are behind or ahead of their weekly step count or MVPA targets. |
| 2.3 Self-monitoring of behaviour | Progress screen | Shows real time progress of weekly step count and MVPA targets. |
| **3. Social support** | | |
| 3.1 Social support (unspecified) | Virtual coach | Encouraging messages to increase physical activity. |
| **4. Shaping knowledge** | | |
| 4.1 Instruction on how to perform a behaviour | Virtual coach | Behavioural options to reach the weekly targets (e.g. a variety of sports). |
| **5. Natural consequences** | | |
| 5.1 Information about health consequences | Virtual coach | Information on health benefits related to physical activity |
| 5.6 Information about emotional consequences | Virtual coach | Information on emotional benefits related to physical activity. |
| **7. Associations** | | |
| 7.1 Prompts/cues | Virtual coach | Defining environmental stimuli (e.g. when you see the stairs at work, take them; put shoes at the door as a cue to walk). |
| **8. Repetition and substitution** | | |
| 8.2 Behaviour substitution | Virtual coach | Encourages substitute habitual behaviour (e.g. take the stairs rather than elevator; run errand by walking instead of by car) |
| 8.4 Habit reversal | Virtual coach | Encourages to reverse of habitual behaviour (e.g. take the stairs rather than the lift; run an errand by walking instead of by car). |
| 8.7 Graded tasks | Background process | The weekly step count and MVPA targets gradually increase. |
| **9. Comparison of outcomes** | | |
| 9.1 Credible source | Virtual coach | Motivational videos of athletes, scientific experts in the field of sports science or Parkinson's disease. |
| **10. Reward and threat** | | |
| 10.4 Social reward | Virtual coach | Positive reinforcement when weekly targets are met. |
| **12. Antecedents** | | |
| 12.5 Adding objects to the environment | Object | Provision of a wrist-worn activity tracker to facilitate monitoring of physical activity behaviour. |

**Supplementary Table 2.** Behavioural change techniques in the Slow-SPEED app according to the BCT Taxonomy (v1). MVPA = Moderate-to-Vigorous Physical Activity.

##### Masked weekly physical activity targets

We inform the participants that the groups differ in the physical activity tasks assigned, but we do not share specific details about the differences between the groups to maintain blinding. Weekly step count and MVPA targets are shown as percentages rather than absolute numbers. This means all participants aim to reach 100% of their step count and MVPA target each week, though the absolute step count and MVPA minutes required differ between the two study arms. Although weekly goals are expressed as percentages, participants can view the actual number of steps taken and total minutes of MVPA for the current and previous day.

**Personalized physical activity regimen in the Slow-SPEED app**

To ensure continued participant motivation throughout the intervention, weekly step count and MVPA targets are adjusted gradually based on individual performance and ability. Step count and MVPA targets will increase linearly each week by 5% up to a maximum of 50% in the intervention group and by 0.5% up to a maximum of 5% in the control group. Beyond this point, targets are personalized based on weekly performance. If a target is met, it continues to increase by 5% or 0.5%. After a target is missed for one week, the target still increases by 5% or 0.5%. Following a second consecutive week in which the target is missed, the target remains unchanged (+0%). If the target is missed for three weeks in a row, the target decreases by 5% or 0.5%, returning to the target that was last met. Each additional failure results in a further reduction of 5% or 0.5%. A successful week resets the sequence and resumes the increase of +5% or 0.5%. This system applies independently to step count and MVPA. Targets will not fall below a 50% increase (intervention) or 5% (control), nor exceed 100% and 10%, respectively. Supplementary Table 3 and Supplementary Figure 1 present a summary and an example of gradual increase of physical activity targets.

| | **Intervention arm** | **Active control arm** |
|---|---|---|
| **Step count and MVPA target increase** | Up to 100% | Up to 10% |
| **Gradual increase (0-50%)** | 5% per week | 0.5% per week |
| **Personalized adjustments (50-100%)** | | |
| Completed week target | + 5% | + 0.5% |
| 1 failed week | + 5% | + 0.5% |
| 2 consecutive failed weeks | + 0% | + 0% |
| 3 consecutive failed weeks | - 5% | - 0.5% |
| **Maximum step count/day** | 20,000 in Slow-SPEED-NL<br>14,000 in Slow-SPEED-UK<br>20,000 in Slow-SPEED-US | 11,000 in Slow-SPEED-NL<br>7,700 in Slow-SPEED-UK<br>11,000 in Slow-SPEED-US |
| **Maximum MVPA minutes/week** | 300 min | 300 min |

**Supplementary Table 3.** Summary of physical activity regimen in the Slow-SPEED app. The 100% and 10% increase is based on the participants' baseline physical activity level during the 4-week eligibility period. MVPA = moderate-to-vigorous physical activity; BCT = behaviour change technique; PD = Parkinson's disease; NL = Netherlands; UK = United Kingdom; US = United States.

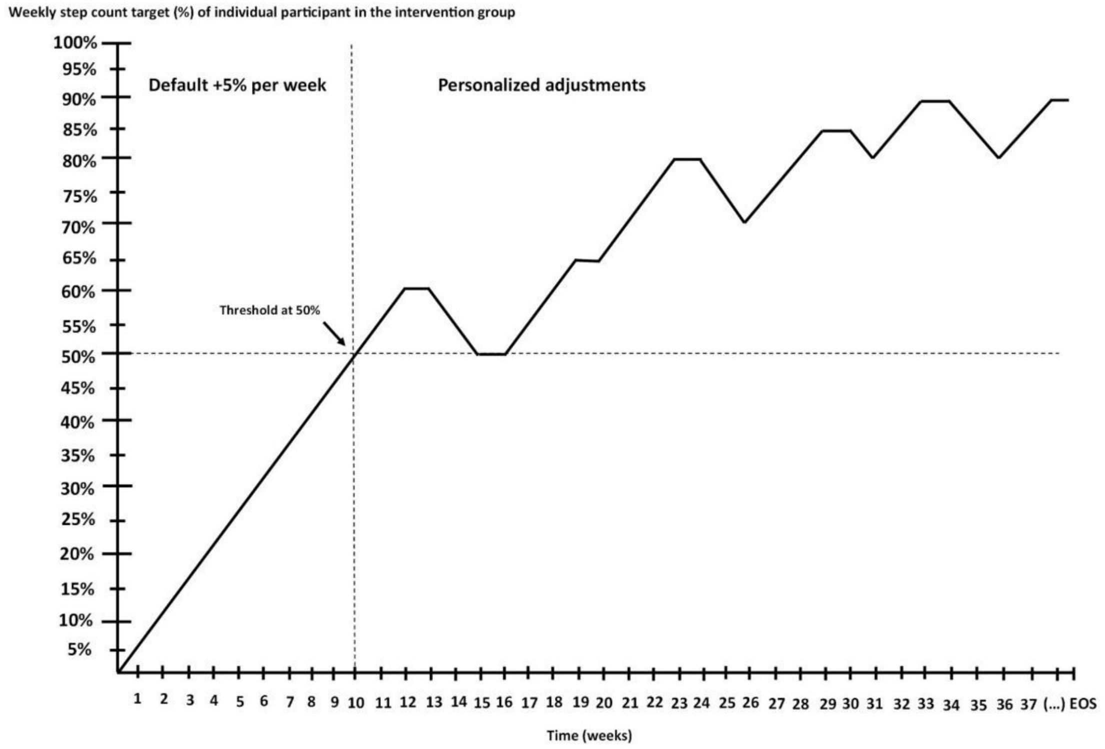

**Supplementary Figure 1**. Hypothetical example of progress of weekly step count targets for a participant in the intervention group. EOS = end of study.

##### Track total progress of physical activity throughout the study

Total progress of absolute step count throughout the study is shown as a virtual journey: participants in Slow-SPEED-NL and Slow-SPEED-UK virtually walk through Europe, while Slow-SPEED-US participants walk from the West Coast of the US to New York. They are also provided a screen with an overview of all weeks to track by percentage whether they have achieved their weekly targets throughout the study duration.

### Supplement 5 – Secondary outcome measures

#### Blood and skin punch sample collection

##### Plans for collection, laboratory evaluation and storage of biological specimens for genetic or molecular analysis in this trial or for future use.

Blood samples are collected within the Slow-SPEED trials for exploratory biomarker research. In Slow-SPEED-NL, blood samples are collected as part of the core study procedures and stored pending completion of the study, with remaining samples stored in a Biobank for future analyses. In Slow-SPEED-UK, blood sampling is optional and limited to a voluntary subgroup of participants, with samples processed and stored in accordance with local laboratory and governance procedures. Supplementary Table 4 shows an overview of the biomarkers of interest. This is subject to change as scientific insights evolve over the course of the trial. Supplementary Table 5 depicts an overview of the sample tubes and processing protocols.

As with blood samples, for Slow-SPEED-UK, skin punch biopsies are optional and used for exploratory biomarker analyses, contributing feasibility outcomes (participant uptake, tolerability, and procedural logistics) and descriptive laboratory outputs related to peripheral α-synuclein pathology, small-fiber nerve integrity, and inflammatory or ageing-related molecular features, without diagnostic, predictive, or disease-modifying intent. Skin punch biopsies will be optionally performed at baseline and end of trial, will be performed under local anaesthesia using a sterile 3-mm punch biopsy instrument, following established research protocols for peripheral tissue sampling in Parkinson's disease and prodromal populations[18]. After skin cleansing with antiseptic solution, 1% lidocaine with epinephrine will be infiltrated subdermally. A single biopsy will be obtained from the cervical paravertebral region (approximately C7-C8, within 3-5 cm of the midline) using firm downward pressure and rotation of the punch to a depth of approximately 3 mm. The tissue sample will be gently removed using sterile forceps, placed immediately into a pre-labelled cryovial, and snap-frozen at -80 °C. Samples will be stored frozen and transported on dry ice for later research analyses. Standard wound care will be applied, and participants will receive post-procedure aftercare instructions.

| **Domain** | **Blood biomarker(s) of interest** |
|---|---|
| Metabolism | Glucose, HbA1c |
| Inflammation | TNF- α[19,20], IL-6[21-24], IL-18[25], IGF-1[26-30], clusterin[31-35], IL-10[20,36,37], PGC-α[38] and irisin[39] |
| Growth factors | BDNF[40-42], GDNF[43], PDGF[44,45], GDF15[46,47], EGF[30,48] |
| Anti-aging marker | Klotho[21,49-51] |
| Pathological protein | α-synuclein[52-54] |
| Neurodegeneration | NfL[55-58] |
| Genetic analyses | LRRK2 G2019S; GBA1 N370S[5] |

**Supplementary Table 4**. List of blood biomarkers. This list may be modified as scientific insights evolve over the course of the trial. HbA1c = hemoglobin A1c; TNF-α = tumor necrosis factor alpha; IL = interleukin; IGF = insulin-like growth factor; PGC-α = Peroxisome proliferator-activated receptor-gamma coactivator alpha; BDNF = brain-derived neurotrophic factor; GDNF = glial cell line-derived; PDNF = platelet-derived neurotrophic factor; GDF15 = growth/differentiation factor 15; EGF = epidermal growth factor; NfL =neurofilament light protein.

| Parameter | Tube 1: Serum (SST) | Tube 2: EDTA-Plasma | Tube 3: cfDNA Plasma | Tube 4: EDTA Whole Blood (DNA storage) | Tube 5: EDTA-Blood (DNA isolation) |
|---|---|---|---|---|---|
| **Tube Type** | SST 8.5 mL with gel (BD 367953) | EDTA K2E 10 mL, no gel, no protease inhibitors (BD 367525) | Streck Cell-free DNA BCT CE, glass (BMD 218996) | EDTA K2E 6 mL, no gel, no protease inhibitors (BD 367864) | EDTA K2E 6 mL, no gel, no protease inhibitors (BD 367864) |
| **N tubes per blood collection** | 2x | 2x | 1x | 1x | 1x, only at beginning of the study. |
| **Volume Collected** | 17 mL | 20 mL | 10 mL | 6 mL | ≥4 mL (6 mL preferred) |
| **Coagulation Time** | 60–120 min at room temperature | Not applicable | Not applicable | Not applicable | Not applicable |
| **Initial Centrifugation** | 2000g, 10 min, room temperature | 2000g, 10 min, room temperature | 2000g, 10 min, room temperature | Not applicable | Not applicable |
| **Second Centrifugation** | Not applicable | Not applicable | 16,000g, 10 min, 4°C | Not applicable | Not applicable |
| **Time Until Freezing / Processing** | Aim: ≤2 hrs; Max: 4 hrs | Aim: ≤2 hrs; Max: 4 hrs | Max: 7 days | Max: 96 hrs | Aim: ≤48 hrs; Max: 1 week |
| **Processing Method** | Clot, centrifuge, aliquot supernatant | Centrifuge, aliquot supernatant | Double centrifugation, aliquot supernatant | Store whole blood tube directly | DNA isolation (Chemagic STAR, Qdoc 084294) |
| **Aliquotation** | 12 aliquots of 500 ul | 12 aliquots of 500 ul | 6 aliquots of 500 ul | No aliquotation; full tube stored | 1 aliquot of 500 ul DNA stock |
| **Aliquot Tube Type** | 2 mL PP screw-cap with O-ring (GREI722301UMC / GREI368380UMC) | 2 mL PP screw-cap with O-ring (GREI722301UMC / GREI368380UMC) | 5 mL PP screw-cap (Fisherbrand 10-500-27) | Original EDTA tube | Matrix tube (2D barcoded, ThermoFisher) |
| **Storage Temperature** | –80°C | –80°C | –80°C | –80°C | –20°C |

**Supplementary Table 5**. Overview of Sample Tubes and Processing Protocols. BCT = blood collection tube; BD = Becton, Dickinson and Company; EDTA K2E = dipotassium ethylenediaminetetraacetic acid; PP = polypropylene; SST = serum separator tube.

#### MRI Acquisition Parameters by Sequence Type

| Parameter | T1-weighted[59] | Resting-State BOLD fMRI[60,61] | Task fMRI[61] | Neuromelanin-sensitive TSE[62-64] | Diffusion Weighted Imaging[65] | R2*, QSM[66] | T2-weighted[67] | FLAIR[68] |
|---|---|---|---|---|---|---|---|---|
| **Target feature** | Global brain atrophy | Cerebral plasticity | Cerebral plasticity | Neurodegeneration | Neurodegeneration | Neurodegeneration | Global brain atrophy | Vascular damage |
| **Region(s) of interest** | Gray and white matter | Putamen and cortex | Putamen and cortex | Substantia nigra, Locus coeruleus | Substantia nigra | Substantia nigra and cortex | Gray and white matter | White matter |
| **Metric** | Tissue volume | Functional connectivity | BOLD-activation | Tissue integrity, microstructure | Tissue integrity, microstructure | Magnetic properties, iron deposition | Tissue volume | Hyperintensity burden |
| **Sequence type** | MPRAGE | Multi-band Multi-echo EPI | Multi-band Multi-echo EPI | T1-weighted TSE | EPI | 3D Multi-echo GRE (FLASH) | 3D TSE | 3D FLAIR |
| **TR (ms)** | 2000 | 735 | 1000 | 890 | 3000 | 40 | 3200 | 5000 |
| **TE (ms)** | 2.03 | 39 | 34 | 13 | 74.40 | 3.87 / 7.83 / 11.79 / 15.75 / 19.71 / 23.67 / 27.63 / 31.59 / 35.53 | 566 | 397 |
| **TI (ms)** | 880 | – | - | – | – | – | – | 1800 |
| **Flip angle (°)** | 8 | 52 | 60 | 120 | 90 | 15 | Variable (T2) | Variable (T2) |
| **Voxel size (mm³)** | 1.0 × 1.0 × 1.0 | 2.4 × 2.4 × 2.4 | 2.0 x 2.0 x 2.0 | 0.2 × 0.2 × 3.0 | 2.0 × 2.0 × 2.0 | 0.8 x 0.8 x 0.9 | 1.0 × 1.0 × 1.0 | 0.5 x 0.5 x 1.0 |
| **FOV (mm)** | 256 | 210 | 210 | 220 | 210 | 217 | 256 | 256 |
| **Acceleration / MB factor** | – | 8 | 6 | – | 3 | 4 | 2 | 3 |
| **Diffusion directions** | – | – | - | – | 104 | – | – | – |
| **B-values (s/mm²)** | – | – | - | – | 0 and 2000 | – | – | – |
| **Volumes acquired** | – | 400 | 1000 | – | – | – | – | – |
| **Phase Encoding Direction** | A >> P | A >> P | A >> P | R >> L | A >> P | R >> L | A >> P | A >> P |
| **Acquisition time** | 4 min 56 s | 10 min 01 s | 10 min | 6 min 48 s | DWI: 5 min 30 s<br>DWI inverted: 18 s | 8 min 47 s | 4 min 40 s | 5 min 17 s |

**Supplementary Table 6**. Overview MRI acquisition parameters by sequence type for the Netherlands trial. All subjects will be scanned on a SIEMENS Prisma 3 Tesla MRI system. fMRI = functional MRI; TSE = Turbo Spin Echo; QSM = Quantitative Susceptibility Mapping; EPI = Echo Planar Imaging; FOV = Field Of View; MB = Multi-band.

#### Axivity AX6 Accelerometer and Gyroscope

In Slow-SPEED-NL and Slow-SPEED-UK, accelerometer and gyroscope data will be collected through passive monitoring using the Axivity AX6 device (Axivity, Ltd., Newcastle upon Tyne, UK). Participants will wear the device for seven consecutive days once per year across the study period. Data acquisition will follow the protocol previously described[69,70]. The sensor placement and settings are shown in Supplementary Figure 2. A separate publication will detail the analysis plan for these sensor data. Sensors are worn simultaneously on the wrist and lower back, while the activity tracker is worn on the opposite wrist.

Predefined analyses will involve the derivation of digital biomarkers for PD motor signs, including gait pattern changes and tremor, using the ParaDigMa toolbox[71]. This toolbox provides validated, open-source algorithms to quantify tremor and arm swing during gait, based on continuous wrist accelerometer and gyroscope data[72,73]. A separate publication will provide more details about the analysis plan for these sensor data

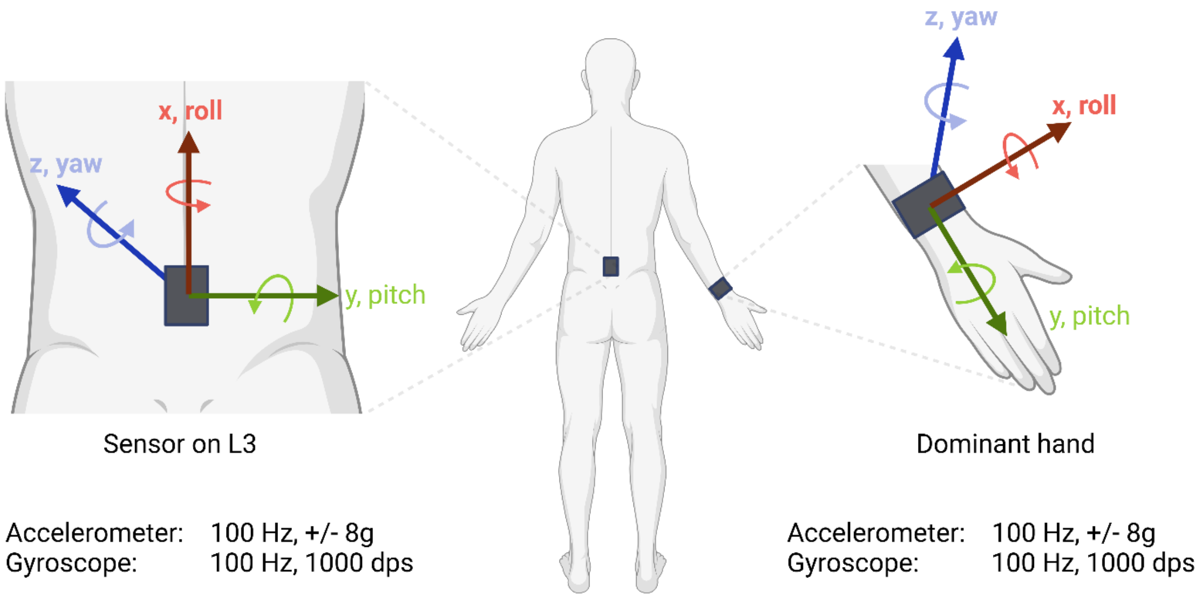

**Supplementary Figure 2.** Axivity AX6 sensor placement. The wrist-worn sensor is worn on the dominant hand (right or left).

#### Supplement 6 – Sample size calculation

**Sample size for Slow-SPEED-NL and Slow-SPEED-UK**

Slow-SPEED-NL and Slow-SPEED-UK are designed as a feasibility study. The primary objectives are to assess feasibility, acceptability, safety, and variability of change in daily step count in adults with iRBD (Slow-SPEED-NL) and OD (Slow-SPEED-UK). The sample size of 110 for Slow-SPEED-NL and Slow-SPEED-UK was selected to provide approximately 80% power – within each population (NL: iRBD, UK: hyposmia defined by UPSIT) – for a treatment difference in sustained increase of daily step counts of 1500 steps per day assuming a standard deviation of 2500 steps per day, 20% loss to follow-up, and a two-tailed 5% type 1 error rate based on a simple two-group t-test. A difference of 1700 steps per day was observed for the increase in daily step count in our STEPWISE pilot trial[74] after 4 weeks between manifest PD patients randomized to 10% vs. 100% targets. Anchored to disease-related patient-reported outcomes of manifest PD patients, 1700 steps per day was identified as a minimum clinically important difference[75]. Person-to-person variation in change in daily step counts from a meta-analysis[76] ranged from a standard deviation of 1600 to 4500 among six randomized trials with a median of 2700, a weighted average of 3100, and a standard deviation of 2000 steps per day in the single trial with at least 1 year follow-up. Power will be greater in the planned mixed-effect model relative to an unadjusted t-test, but the ultimate achieved power of Slow-SPEED-NL and Slow-SPEED-UK is unknown given uncertainties in the magnitude of any sustained increase in daily step count and of person-to-person variation in step count change over 24 months (Slow-SPEED-NL) or 18 months (Slow-SPEED-UK). The matched sample size of Slow-SPEED-NL and Slow-SPEED-UK support harmonisation for pooled data analyses.

**Sample size for Slow-SPEED-US**

The sample size of 600 for Slow-SPEED-US was selected to provide approximately 80% power for a treatment difference in sustained increase of daily step counts of 1500 steps per day within each of four strata (LRRK2 or GBA genetic status and female or male sex) assuming a standard deviation of 2500 steps per day, 40% loss to follow-up (higher than the drop-out rate assumed in Slow-SPEED-NL and Slow-SPEED-UK), and a two-tailed 5% type 1 error rate based on a simple two-group t-test. Assuming consistent increases across the strata, power for the full cohort will be >99% for this co-primary endpoint. Given that the prodromal load score is still under development, we do not have preliminary data from which to estimate an effect of exercise or person-to-person change in prodromal load scores over 36 months. For the full cohort, we would have >90% power if the effect size for change in prodromal load score were at least 0.34, yielding at least 80% power for observing a benefit on both co-primary endpoints.

**Recruitment strategies**

Each trial has a main source of participants in each country. In Slow-SPEED-NL, participants are identified via Dutch sleep clinics (Sleep Wake Center, Stichting Epilepsie Instellingen Nederland (SEIN) and Sleep Medicine Center Kempenhaeghe) and other sleep-accredited medical centres across the country and referred to Radboudumc for enrolment and study procedures. In Slow-SPEED-UK participants are identified from PREDICT-PD, a pioneering, online, longitudinal community-based study that has already enrolled thousands of adults across the UK to identify risk factors for PD[15], as well as specialist Ear, Nose and Throat clinics for olfactory disorders at the James Paget University Hospitals National Health Service Foundation Trust. Participants are referred to

Queen Mary University of London for enrolment and study procedures. In Slow-SPEED-US participants are identified via the research participant base of the 23andMe Research Institute, which provides direct-to-consumer genetic testing for opt-in participation in research studies. Slow-SPEED-US participant are referred to the University of Rochester for enrolment and study procedures. We also advertise the trials on social media (LinkedIn, X, and Instagram) and provide study-related information through Parkinson's disease-related channels specific to each country.

#### Supplement 7 – Treatment assignment, blinding and participant retention

##### Treatment assignment and randomisation

Participants are randomised in a 1:1 ratio to the intervention or active-control group using a computer-generated randomisation tool in the Castor EDC data management system[77] in Slow-SPEED-NL and Slow-SPEED-US and REDCap randomisation tool in Slow-SPEED-UK[78] .
The randomisation schedules are slightly different between the three trials as the targeted sample sizes and stratification differ. Slow-SPEED-NL includes random permuted blocks (block sizes: 2 and 4) stratified by sex (male, female). Slow-SPEED-UK includes random permuted blocks (block sizes: 2, 4 and 6) stratified by age (40-59 vs ≥60 years), baseline activity (<5,000 vs 5,000-6,999 steps/day), and hyposmia severity (UPSIT). Slow-SPEED-US includes random permuted blocks (block sizes: 4 and 6) stratified by sex (male, female) and risk variant (*LRRK2*, *GBA1*).

##### Blinding of participants

Treatment assignment is performed by researchers who are not involved in the administration of the intervention or data collection. Group allocation is manually entered on the server side of the Slow-SPEED app based on the randomisation. Group allocation is concealed from all other members of the study team as well as from participants. After group allocation, participants are given full access to the app. The app is visually identical for participants in both groups and include masked weekly physical activity targets to enhance blinding. Blinding of participants is checked at the end of all three trials by asking participants whether they think they were randomised in the group with a smaller or larger increase in physical activity.
Participants will not be unblinded prior to data lock unless there is a health-related concern for the participant, in the opinion of the investigator, or upon request from an accredited medical research ethics committee (MREC). Any emergency unblinding will be authorised by the principal investigator and fully documented.

##### Criteria for discontinuing or modifying allocated interventions

Participants may withdraw consent and discontinue their participation at any time, without providing a reason and without any impact on their future medical care. Participants who choose to withdraw from the study are invited to complete follow-up assessments. In all cases, participant safety, autonomy, and confidentiality are prioritized in accordance with the Declaration of Helsinki (2024), the General Data Protection Regulation (GDPR) in Europe, and applicable US regulations.

##### Strategies to improve adherence to interventions and promote retention

The Slow-SPEED app is the main strategy for adherence to the intervention, providing participants with structured guidance and motivation. The study team will offer technical support for any issues with the apps or devices used in this study. To foster continued engagement, participants will receive regular trial updates and newsletters, as well as invitations to attend virtual public engagement events. In addition, in Slow-SPEED-UK, all participants are offered complimentary gym membership and access to Ordnance Survey maps for walking routes as engagement and retention incentives and are not part of the Slow-SPEED intervention. Uptake of these resources is monitored optionally via self-report questionnaires.

##### Relevant concomitant care permitted or prohibited during the trial

We allow participants to continue all usual medical care. To ensure we can accurately measure the effect of the Slow-SPEED intervention, we ask participants to inform us if they join other exercise or physical activity promotion trials during follow-up. Participation in observational studies or other trials is permitted. If participants choose to take part in other lifestyle programmes or exercise classes, this will not be prohibited, but the trial name is recorded and accounted for during analyses.

#### Supplement 8 – Oversight and Monitoring

##### Composition of the coordinating centre and trial steering committee

Slow-SPEED-NL and Slow-SPEED-US are coordinated by the research team of Radboudumc in the Netherlands. Communication with Slow-SPEED-US participants is supported by the Center for Health and Technology of the University of Rochester, USA. Slow-SPEED-NL and Slow-SPEED-US are monitored by an independent monitor from the Radboudumc according to the guidelines of the 'Nederlandse Federate van Universitair medisch centra'. In Slow-SPEED-UK, sponsorship and governance are provided by QMUL through the Joint Research Management Office (JRMO). Ethics approval is obtained from the Health Research Authority (HRA) and an NHS Research Ethics Committee (REC). The trial runs under UK Good Clinical Practice (GCP), and the UK Policy Framework for Health and Social Care Research. An independent monitor or auditor appointed under JRMO procedures oversees compliance and data quality, with serious breaches and required safety reports notified to the HRA and REC.

##### Adverse event reporting and harms

All self-reported adverse events and serious adverse events (SAE) are recorded. In Slow-SPEED-NL and Slow-SPEED-UK, participants are requested to provide spontaneous reports of any injuries or health changes, whereas in Slow-SPEED-US, this information is collected through active solicitation using scheduled reminders. SAEs that result in death or are life-threatening will be reported to the accredited medical ethics committee that approved the protocol within 7 days of the researchers' first awareness of the event. This is followed by a period of up to 8 days to complete the initial preliminary report. All other SAEs will be reported within 15 days (or 10 business days) after the researchers' first knowledge of the SAE. No compensation will be provided for any harm resulting from trial participation. If medical attention is required, participants will be referred to their own healthcare provider for evaluation and treatment.

**Plans for communicating important protocol amendments to relevant parties**
Amendments to the research protocol may be introduced in the future if new scientific insights provide a clear rationale for changes. Substantial amendments will be implemented only after approval by the applicable medical ethics committee. Participants will be informed when substantial amendments are made to the participant information sheet or the informed consent. Non-substantial amendments will not be submitted to the medical ethics committee or competent authority but will be documented, recorded and filed by the sponsor.

#### Supplement 9 – Data collection and management

##### Data management
Participants are provided a pseudonymized Google account to connect their own smartphone to the activity tracker. The data collected via the activity tracker are stored on a server hosted by Rootnet B.V. (Nijmegen, The Netherlands). The server is protected by software updates, daily backups and 24/7 monitoring conforming to ISO27001 and NEN 7510. Research employees from Radboudumc, University of Rochester, and Queen Mary University of London (QMUL) can access these data through an institutional login-linked, username and password-protected server of the app. In parallel, Google stores all data collected on Fitbit activity trackers on their own servers and no personal identifiers are used due to use of pseudonymized Google accounts. Participants are informed about this in the study information and informed consent form. Secondary outcome measures collected in-clinic (blood pressure, orthostatic hypotension) and at home (questionnaires, UPSIT olfactory test) are directly entered into secured and certified data management systems (Castor EDC for Slow-SPEED-NL and Slow-SPEED-US and REDCap for Slow-SPEED-UK). Both data management systems provide an audit trail. For Slow-SPEED-UK, REDCap is hosted on secure QMUL servers and managed in accordance with UK Data Protection Act 2018, UK GDPR, and Health Research Authority guidance. Access is role-based and restricted to authorised study personnel.

The data from the Axivity sensors are manually extracted by research staff via a wired connection and subsequently stored on a secured department server of Radboudumc for Slow-SPEED-NL and on secure QMUL servers for Slow-SPEED-UK . Data collected from MRI-scans are stored at the Donders Institute infrastructure and archived on the Donders repository. Blood tubes are identified with a random 7-digit code and stored upon completion of the trial. The samples are stored at the Radboudumc Biobank for Slow-SPEED-NL and at Wolfson Laboratory G016, Wolfson Institute of Population Health for Slow-SPEED-UK. Participants are provided with an anonymized account to use the Roche PD research app on their own phone. Data from this app is temporarily stored locally on the smartphone, after which is encrypted, transferred to, and stored on secured servers of Hoffmann – La Roche Ltd (Basel, Switzerland). Once transferred, collected data is deleted from the smartphone. Anonymized data from the Roche app is transferred to the study team every nine months, after which this is stored on the secured department server of Radboudumc (Slow-SPEED-NL and Slow-SPEED-US) or QMUL (Slow-SPEED-UK). Participants are informed in the study information and informed consent about Hoffmann – La Roche Ltd receiving these data.

**Confidentiality**
All personal data are handled in compliance with the EU General Data Protection Regulation. For Slow-SPEED-NL and Slow-SPEED-US, personal information are kept in our participant registration system Salesforce (Salesforce Inc, San Francisco, USA) which provides an audit trail. Personal data from participants in Slow-SPEED-US are stored on Microsoft Teams in parallel (Microsoft Teams, Microsoft Corp., Redmond, WA, USA). For Slow-SPEEP-UK, personal information are stored in encrypted files at protected department servers at QMUL, accessible only to authorised study staff. Participants are assigned a unique personal identification code consisting of a combination of three random capital letters and three digits (e.g., DGH398) for data collection through the Slow-SPEED app and for MRI-scans. A random 10-digit code is constructed for the Roche PD research app and a random 7-digit code is used for the blood samples. Personal data are kept separately from the experimental data. Data stored in Castor EDC or REDCap are username and password protected with access provided only to dedicated research staff. Data will be locked and stored for at least 15 years after the study ends.

#### Supplement 10 – Statistical analysis of secondary outcomes

*Feasibility*
The secondary outcomes for assessing feasibility are within-subject change in the number of minutes performing moderate-to-vigorous physical activity (MVPA). We will calculate for each participant the number of minutes performing MVPA and physical fitness biomarkers per four-week intervals from the four weeks prior to baseline through the 104-week treatment period in Slow-SPEED-NL (total 27 time points), 78-week period in Slow-SPEED-UK (total 20 time points) and 156-week treatment period in Slow-SPEED-US (total 40 time points). The same model used for the primary outcome will be used for MVPA. As age-adjusted HRmax may be biased in participants on rate-limiting/chronotropic medications or with autonomic dysfunction, we will adjust the model for their presence, and run sensitivity analyses using heart-rate–independent intensity proxies (e.g., cadence-based thresholds) to corroborate MVPA findings.

Secondary outcomes related to the feasibility of remotely monitoring the intervention, as well assessments of the usability and evaluation of the Slow-SPEED app and barriers and motivators to engage in physical activity will be reported descriptively and will not undergo statistical analysis.

*Preliminary efficacy*
The secondary study outcomes to assess preliminary efficacy are within-subject change in digital biomarkers for physical fitness, motor symptoms, non-motor symptoms, blood-based biomarkers, brain imaging biomarkers, quality of life, and functional status (see Table 3 for the frequency of each assessment). The same model used for the primary outcome will be used for these measures with the number of levels of the time variable reduce to reflect the planned follow-up schedule of each secondary outcome.

Brain-imaging markers will be analysed differently for each sequence used:
a. T1, T2, FLAIR: The difference of region-specific white and gray matter, T1/T2 ratio of the basal ganglia and white matter hyperintensities on baseline MRI and follow-up MRI will be calculated per

individual. The resulting delta will be subjected to a linear regression testing effect of the group (intervention vs active control) on change over time. Analysis will be done separately for each variable (3 total).
b. Resting-state functional MRI: Functional coupling between striatal subregions and the cerebral cortex will be analysed using seed-to-voxel and node-based functional connectivity analyses on individual and group-level using the General Linear Model (GLM) on resting state functional MRI data[60,79].
c. Task-related functional MRI: Task-related cerebral activity will be estimated and compared between groups using the GLM in Statistical Parametric Mapping (SPM12; http://www.fil.ion.ucl.ac.uk/spm). We will focus on regions of interest: the putamen and the dorsal premotor and posterior parietal cortex (motor task), based on previous work using the same task in PD[79,80]. As outlined below, task-related activity in these brain regions will be compared between groups (to test for exercise-specific effects) and time points (to test for longitudinal changes). Specifically, we will perform a repeated-measures ANOVA with between-subjects factor GROUP (i.e. the intervention group versus active controls), and within-subjects factors TIME (baseline versus two year or three years follow-up) and condition (CHOICE: one-choice versus multiple-choice).
d. Diffusion weighted imaging: Change in substantia nigra free water between baseline MRI and follow-up MRI will be calculated per individual, as done before[80]. The resulting delta will be subjected to a linear regression testing effects of GROUP (intervention vs active control) on change over time.
e. Neuromelanin MRI: Change in substantia nigra intensity and volume and locus coeruleus intensity between baseline MRI and follow-up MRI will be calculated per individual and corrected for signal intensity in a control region in the brain stem, as done before[81,82]. The resulting delta will be subjected to a linear regression testing effects of GROUP (intervention vs active control) on change over time.
f. R2*/QSM: Change in QSM signal in the basal ganglia and substantia nigra between baseline MRI and follow-up MRI will be calculated per individual, as done before[83]. In addition, we will also quantify QSM across cortical and subcortical areas, both in grey matter and superficial white matter. The resulting delta will be subjected to a linear regression testing effect of GROUP (intervention vs active control) on change over time.

As part of the exploratory analyses, MRI outcome measures may be analyzed using the same statistical model as specified for the primary objective, provided the outcomes are continuous in nature and suitable for this modeling approach. Subgroup analyses will be conducted to assess the effect of treatment assignment on the outcomes as mentioned before, with adjustment for age, sex, socio-economic status or comorbidity. In the US, subgroup analyses will be conducted to assess the effect of treatment allocation adjusted for genetic variant (LRRK2, GBA1). We might conduct exploratory analyses in the future, if new insights are published during the duration of this study.

**Other study parameters**
Baseline characteristics will be included in a descriptive way.

**Interim analyses**

Interim analyses will be performed to evaluate futility. The timing of the interim analyses and the criteria used for a non-binding recommendation to stop a given Slow-SPEED trial early are listed in the table below.

| Study attribute | Slow-SPEED-NL | Slow-SPEED-UK | Slow-SPEED-US |
|---|---|---|---|
| **Participants** | 50 | 50 | 150 total |
| **Duration** | 12 months | 12 months | 12 months |
| **Aimed difference in step count (Intervention vs. Control)** | ≥ 500 steps/day average | ≥ 500 steps/day average | ≥ 500 steps/day average |
| **If step count goal not reached** | Study will be stopped | Study will be stopped | Study will be stopped |

**Supplementary table 7.** Pre-specified interim analyses. The average step count per day in the four weeks prior to baseline will be compared to the last four weeks of the 12-month period.
